# Capability impacts of the Covid-19 lockdown in association with mental well-being, social connections and existing vulnerabilities: an Austrian survey study

**DOI:** 10.1101/2020.11.14.20231142

**Authors:** Judit Simon, Timea M. Helter, Ross G. White, Catharina van der Boor, Agata Łaszewska

## Abstract

**Background:** Impacts of the Covid-19 pandemic and its public health measures go beyond physical and mental health and incorporate wider well-being impacts in terms of what people are free to do or be. We explored these capability impacts of the Covid-19 lockdown in association with people’s mental well-being, social support and existing vulnerabilities in Austria.

**Methods:** Adult Austrian residents (n=560) provided responses to a cross-sectional online survey about their experiences during Covid-19 lockdown (15 March-15 April 2020). Instruments measuring capabilities (OxCAP-MH), depression and anxiety (HADS), social support (MSPSS) and mental well-being (WHO-5) were used in association with six pre-defined vulnerabilities using multivariable linear regression.

**Results:** 31% of the participants reported low mental well-being and only 30% of those with a history of mental health treatment received treatment during lockdown. Past mental health treatment had a significant negative effect across all outcome measures with an associated capability well-being score reduction of -6.54 (95%CI: -9.26,-3.82). Direct Covid-19 experience and being ‘at risk’ due to age and/or physical health conditions were also associated with significant capability deprivations. When adjusted for vulnerabilities, significant capability reductions were observed in association with increased levels of depression (−1.79) and anxiety (−1.50), and significantly higher capability levels (+3.77) were associated with higher levels of social support. Compared to the cohort average, individual capability impacts varied between -9% for those reporting past mental health treatment and +5% for those reporting one score higher on the social support scale.

**Conclusions:** Our study is the first to assess the capability limiting aspects of a lockdown in association with specific vulnerabilities. The negative impacts of the Covid-19 lockdown were strongest for people with a history of mental health treatment. In future public health policies, special attention should be also paid to improving social support levels to increase public resilience.

## Introduction

The recently discovered coronavirus, known as severe acute respiratory syndrome coronavirus 2 (SARS-CoV-2), has spread globally within a span of a few months since December 2019 [1]. The Covid-19 disease caused by the virus was declared as a pandemic by the World Health Organisation (WHO) on 11^th^ March 2020. Initial evidence suggested that the infection has a high effective reproduction rate and older populations and those with underlying health conditions are at high risk of severe disease and death, thereby forcing numerous countries into temporary lockdowns to limit the spread of the disease. Consequently, the Covid-19 pandemic went from a direct health emergency to a systemic crisis affecting people’s lives in multiple ways [2]. The Covid-19 impacts are unprecedented because of its evolution from a health shock to a global economic and social crisis [2].

Substantial evidence from the past studies of the impacts of Severe Acute Respiratory Syndrome, Middle East Respiratory Syndrome, and Ebola epidemics on the suffering individuals and the healthcare providers showed substantial neuropsychiatric linkage [3]. There is an increasing amount of research related to the impacts Covid-19 on people’s mental health and well-being [3-22]. Beside the direct health impact, public health emergencies may also affect individuals and communities through isolation, stigma, job insecurity, or inadequate resources for medical response [15]. These effects generate a range of emotional reactions, and are expected to be particularly prevalent among those individuals who contract the disease, or are at increased risk due to their age or pre-existing medical conditions [15]. Evidence from previous pandemics shows that individuals who contract the disease experience fear, anxiety, emotional distress, and post-trauma stress symptoms [3]. The mental health/well-being impacts of the pandemic can be even more significant for those who are prone to psychological problems [6].

Impacts of this pandemic and its public health measures go beyond physical and mental health and incorporate wider well-being impacts in terms of what people are free to do or be. Due to these complexities, the assessment of personal consequences related to well-being is challenging and may be best addressed within the conceptual framework of the capability approach introduced by Amartya Sen in the early 1980s [23]. The capability approach proposes that well-being is determined by people’s freedom to engage in forms of being and doing that are of intrinsic value to the person [23]. Beside the recently proposed use of the capability framework in the understanding of policy challenges [24], this freedom aspect can be interpreted in the narrower mental health context as both the actual capabilities of a person, for instance, good mental health, and the processes that enable them, for instance, legal regulations [25]. Not only has the Covid-19 outbreak had a profound psychological impact, but it also affects personal freedoms to engage in behaviours that are consistent with subjectively held values, for instance, visiting loved ones, engaging in recreational activities, spending time outdoors. Despite these important links, the connection between pandemics and individual capabilities have not yet been researched.

In Europe, Austria stood out as a nation that adopted aggressive and early strategies and thereby saw a smaller proportion of deaths from Covid-19 compared to some other European countries [26]. The first Covid-19 case in Austria was reported on 25 February 2020 [27]. The Austrian government issued general laws to contain the epidemic by restricting social contacts and imposing strict lockdown measures from 16 March onwards [27] most of which have been lifted gradually since 15 April. Early studies assessing the Covid-19 and related public health measures impacts have found significant impact on the mental health of the Austrian population. The studies found that symptoms of moderate to severe anxiety and depression have tripled in Austria, and 8-13% of the population showed severe depression and 6-11% severe anxiety symptoms [28, 29].

Despite the increasing number of studies exploring the Covid-19 impact on mental health/well-being, information is missing on the broader capability impact of the pandemic. Hence, this study aimed to explore the impact the Covid-19 lockdown period on people’s capabilities in association with mental health/well-being and social support, especially in the case of specific vulnerable groups in Austria. Vulnerable groups were pre-defined: (i) being categorised as ‘at risk’ group based on age and/or pre-existing physical health conditions; (ii) self-reported mental health treatment prior to the coronavirus pandemic; (iii) direct exposure to Covid-19 (having symptoms or being tested positive); (iv) indirect exposure to Covid-19 through a family member/friend; (v) having employment status impacted by the lockdown; or (vi) being categorised as critical worker.

## Methods

### Study design, data collection and participants

Cross-sectional data were collected via an online survey in May/June 2020, with all questions, including standardised outcome instruments, referring to the one-month lockdown period in Austria between 15 March and 15 April 2020.

Participants were recruited using convenience sampling. Adults (≥18 years), with sufficient German knowledge, access to the online survey, and residency in Austria at the time of the Covid-19 outbreak were able to participate. The survey was developed in the SoSci online survey platform (Version 2), which is a publicly available tool and is free of charge for academic research [30]. The weblink of the survey was included in an advert, along with a QR code, that was circulated via social media platforms (including Facebook, Twitter, WhatsApp, etc.) and emails targeting a wide range of individuals and organisations throughout Austria.

Respondents who provided sociodemographic and Covid-19-related information and completed at least one standardised instrument were considered for analysis. Those participants who discontinued the survey before fully completing at least one standardised instrument were excluded from the analyses.

### Survey and instruments

The online survey consisted of the participant information and consent forms followed by a section on sociodemographics. Subsequent sections assessed people’s perceptions about the Covid-19 outbreak and the public health measures in place during the lockdown in Austria in response to the outbreak. The final part of the questionnaire consisted of four self-reported standardised and validated outcome instruments, which were used to assess capability well-being (OxCAP-MH), depression and anxiety levels (HADS), social support (MSPSS) and mental well-being (WHO-5) similar to a parallel linked survey in the UK [31].

The Oxford CAPabilities questionnaire-Mental Health (OxCAP-MH) instrument was developed by Simon et al. in 2013 [32]. It is specifically designed to capture different well-being dimensions within the capability framework in the area of mental health across 16 items. The OxCAP-MH is scored on a 0–100 scale, with higher scores indicating better capabilities. The German version of the OxCAP-MH [33] was obtained from the authors for the study.

The Hospital Anxiety and Depression Scale (HADS) was developed by Zigmond and Snaith in 1983 [34]. The HADS was found to perform well in assessing the presence and severity of anxiety disorders and depression, also beyond the hospital setting, including the general population [35]. The questionnaire is divided into Anxiety (HADS-A) and Depression (HADS-D) subscales both containing seven items scored on a four-point scale from zero (not present) to three (considerable). Both the HADS-A and HADS-D subscales are scored from 0-21, with higher scores indicating higher anxiety or depression levels. Normal, borderline and abnormal anxiety/depression scores are defined as 0-7, 8-10 and 11-21, respectively [34]. The German translation of HADS was obtained from Hogrefe Publishing Group.

The Multidimensional Scale of Perceived Social Support (MSPSS) is a self-reported measure of subjectively assessed social support developed by Zimet et al. in 1988 [36]. The questionnaire can be divided into three subscales, each addressing a different source of support: Family, Friends, and Significant Other. Low, moderate and high support are defined as <3, 3-5 and >5, respectively [36]. An official German translation of MSPSS was obtained from the developer of the original English version.

The World Health Organisation-Five Well-being Index (WHO-5) is a short self-reported measure of current mental well-being introduced in 1998 by the WHO Regional Office in Europe [37]. Respondents are asked to rate how well each of the five statements about positive well-being applied to them in the given period from 5 (all of the time) to 0 (none of the time). The WHO-5 is scored 0-25, with higher scores representing higher well-being [38]. The German translation of the WHO-5 is available in the public domain without registration.

### Statistical analysis

Anonymous data were extracted from the online survey and checked for logical inconsistencies. Six hypothesised associations between higher levels of mental health symptoms and lower levels of well-being were tested for pre-defined vulnerabilities: i) At risk group; ii) Past mental health treatment; iii) Direct Covid-19 experience; iv) Indirect Covid-19 experience; v) Employment status affected by Covid- 19; and vii) Critical worker.

Individuals were defined as ‘at risk’ if they were aged 65 years or over and/or had a self-reported underlying physical health condition including diabetes, heart/cardiovascular disease, stroke/cerebrovascular disease, lung disease, liver disease, or cancer. Participants who reported mental health service use prior to the period of interest were categorised as ‘having past mental health treatment’. Participants with ‘direct Covid-19 experience’ were those who tested positive for Covid-19 or experienced Covid-19 symptoms, but were not tested. ‘Indirect Covid-19 experience’ was defined as having a friend and/or family member infected or knowing someone who died of Covid-19. Participants with ‘employment status affected’ were those who reported losing their job due to the pandemic or being sent to short-time working (German ‘*Kurzarbeit’*). Finally, participants who reported having a job categorised by the government as critical worker, e.g. healthcare staff, police officer or food supply worker, were defined as ‘critical workers’.

Correlations between the different outcome measures were explored using Pearson’s correlations and interpreted as small <0.3, moderate 0.3-0.49, or large ≥0.50 [39]. In order to explore the impacts of the Covid-19 lockdown on capabilities, mental health/well-being and social support in association with pre- defined vulnerabilities, multivariable linear regression analyses were conducted using the OxCAP-MH, HADS-D, HADS-A, MSPSS and WHO-5 scores as dependent variables and binary vulnerable group defining variables as independent variables. Analyses were adjusted for age, gender, having children, education level and initial employment status.

The magnitude of impact of current depression, anxiety and social connections on capabilities was investigated separately in a multivariable regression analysis adjusted for all measured sociodemographic characteristics and pre-defined vulnerabilities. Significance level of p<0.05 was considered in all analyses. Analyses were conducted on complete cases in STATA v.15.1 [40].

## Results

### Participant characteristics

848 persons accessed the survey, out of whom 560 respondents (74.1% female, mean age 40.2 years) completed it and was included in the analyses (Figure 1). The average time needed to complete the survey was 17 minutes.

**Figure 1.**
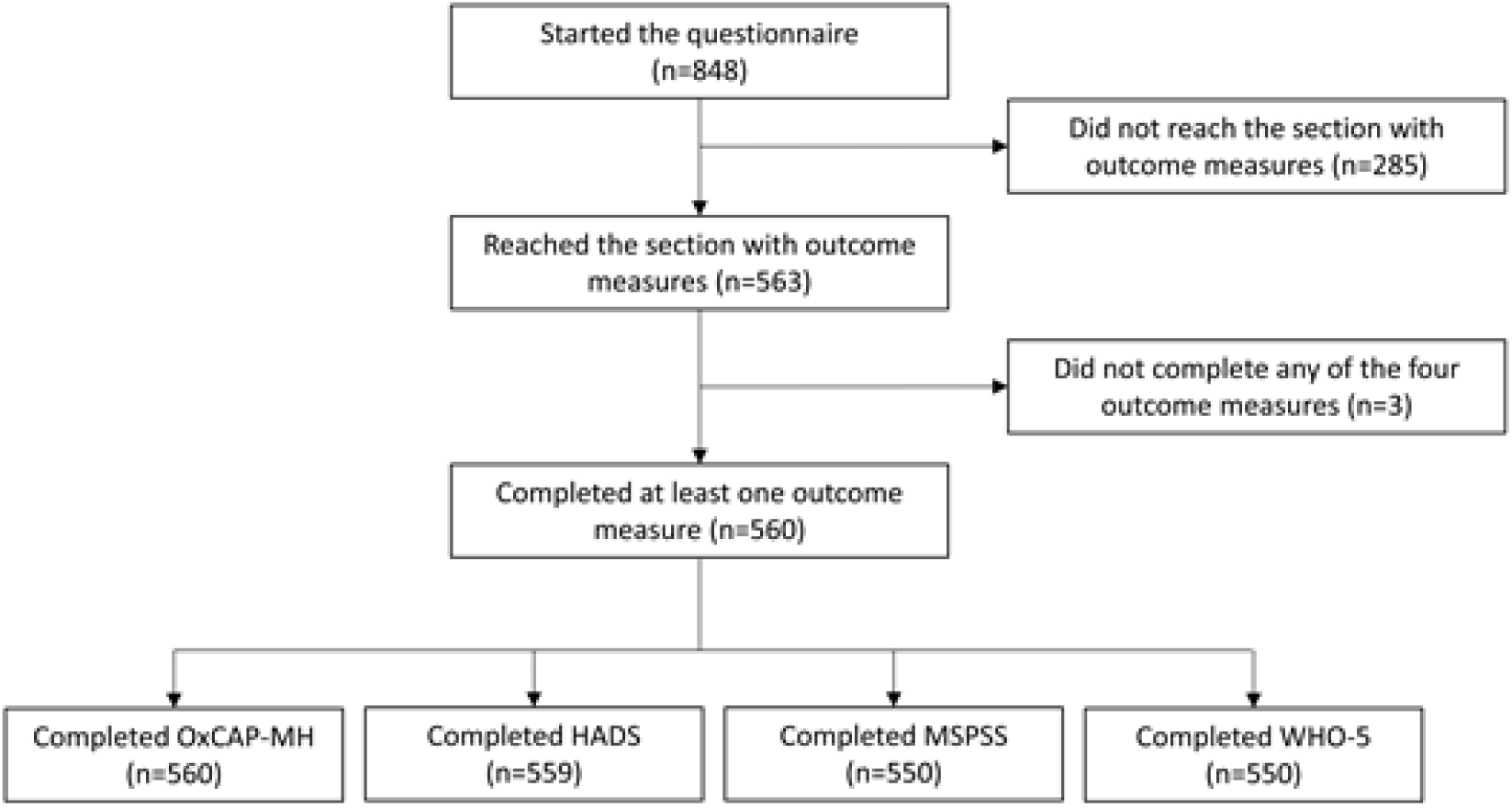
Survey flowchart.

The majority of participants were Austrian citizens (87%) and employed at the beginning of the Covid- 19 lockdown (73%). More than half of the survey participants (56%) had children, 52% were married or had a registered partnership. Full data on sociodemographic characteristics in comparison to the official Austrian population statistics, with respect to age, gender, distribution of population across federal states [41], migration background [42], education level [43], and employment status [44], are shown in Table 1.

**Table 1.**
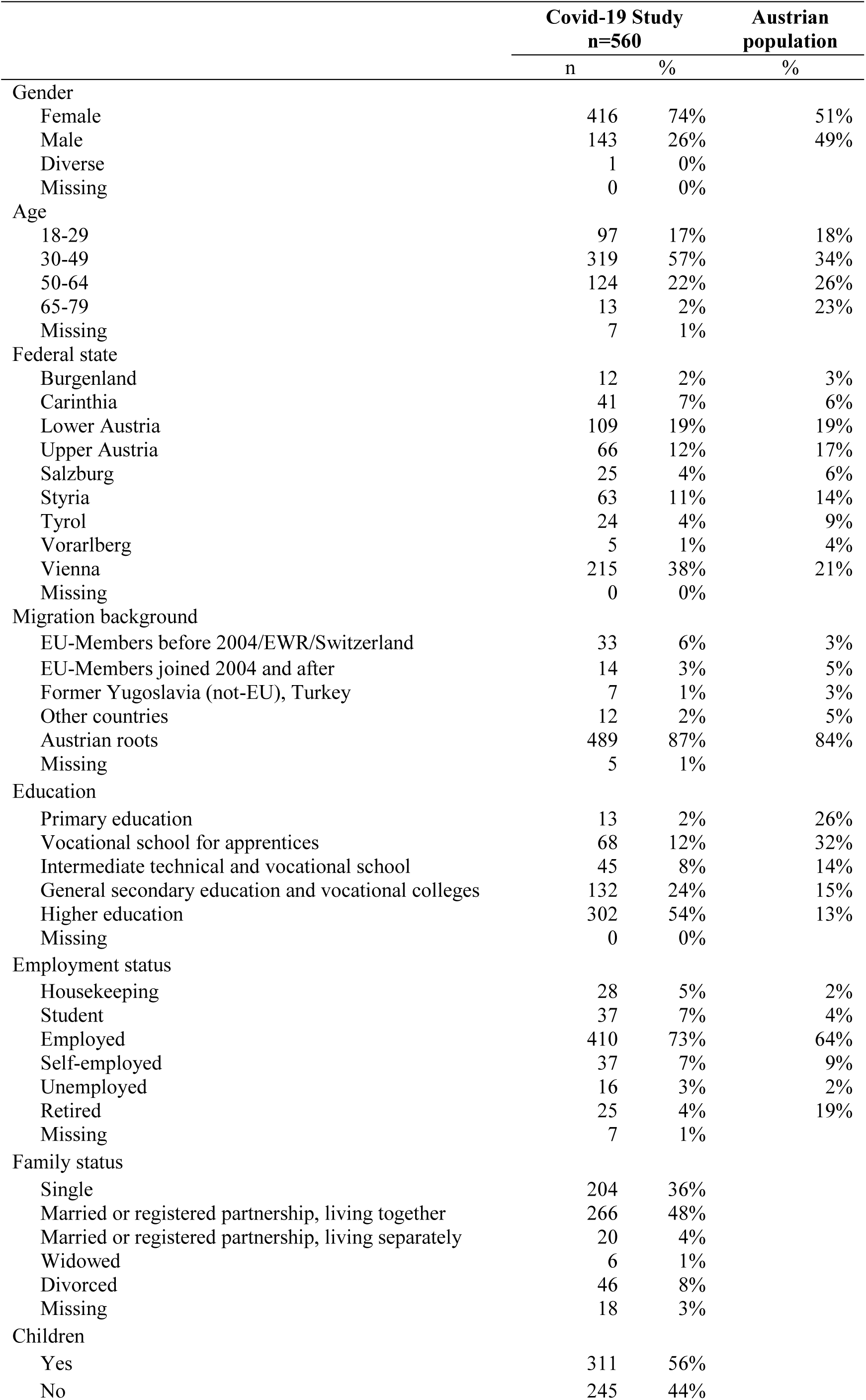
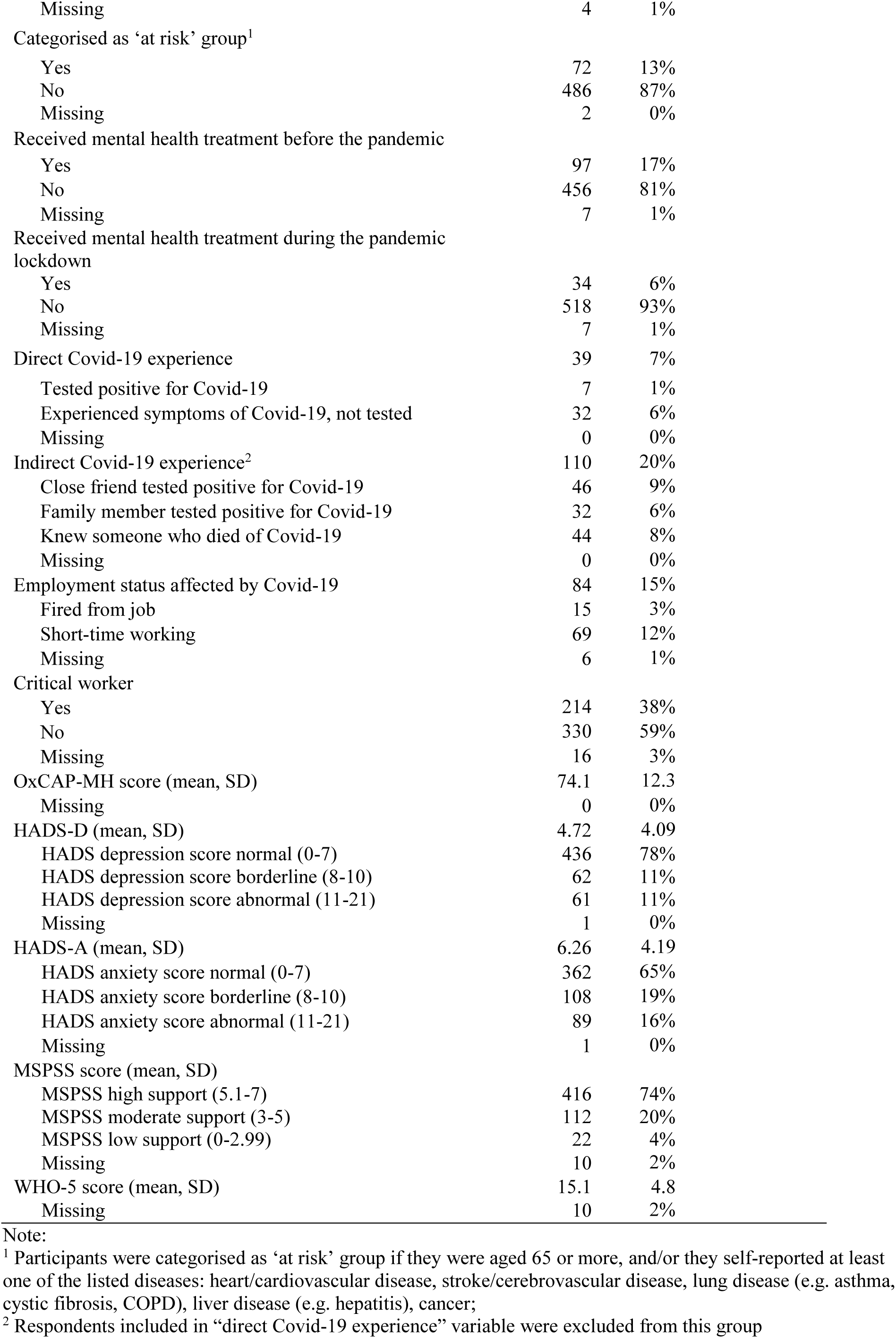
Characteristics of the survey cohort

|  | Covid-19 Study<br>n=560 |  | Austrian<br>population |
| --- | --- | --- | --- |
|  | n | % | % |
| Gender |  |  |  |
| Female | 416 | 74% | 51% |
| Male | 143 | 26% | 49% |
| Diverse | 1 | 0% |  |
| Missing | 0 | 0% |  |
| Age |  |  |  |
| 18-29 | 97 | 17% | 18% |
| 30-49 | 319 | 57% | 34% |
| 50-64 | 124 | 22% | 26% |
| 65-79 | 13 | 2% | 23% |
| Missing | 7 | 1% |  |
| Federal state |  |  |  |
| Burgenland | 12 | 2% | 3% |
| Carinthia | 41 | 7% | 6% |
| Lower Austria | 109 | 19% | 19% |
| Upper Austria | 66 | 12% | 17% |
| Salzburg | 25 | 4% | 6% |
| Styria | 63 | 11% | 14% |
| Tyrol | 24 | 4% | 9% |
| Vorarlberg | 5 | 1% | 4% |
| Vienna | 215 | 38% | 21% |
| Missing | 0 | 0% |  |
| Migration background |  |  |  |
| EU-Members before 2004/EWR/Switzerland | 33 | 6% | 3% |
| EU-Members joined 2004 and after | 14 | 3% | 5% |
| Former Yugoslavia (not-EU), Turkey | 7 | 1% | 3% |
| Other countries | 12 | 2% | 5% |
| Austrian roots | 489 | 87% | 84% |
| Missing | 5 | 1% |  |
| Education |  |  |  |
| Primary education | 13 | 2% | 26% |
| Vocational school for apprentices | 68 | 12% | 32% |
| Intermediate technical and vocational school | 45 | 8% | 14% |
| General secondary education and vocational colleges | 132 | 24% | 15% |
| Higher education | 302 | 54% | 13% |
| Missing | 0 | 0% |  |
| Employment status |  |  |  |
| Housekeeping | 28 | 5% | 2% |
| Student | 37 | 7% | 4% |
| Employed | 410 | 73% | 64% |
| Self-employed | 37 | 7% | 9% |
| Unemployed | 16 | 3% | 2% |
| Retired | 25 | 4% | 19% |
| Missing | 7 | 1% |  |
| Family status |  |  |  |
| Single | 204 | 36% |  |
| Married or registered partnership, living together | 266 | 48% |  |
| Married or registered partnership, living separately | 20 | 4% |  |
| Widowed | 6 | 1% |  |
| Divorced | 46 | 8% |  |
| Missing | 18 | 3% |  |
| Children |  |  |  |
| Yes | 311 | 56% |  |
| No | 245 | 44% |  |

|  |  |  |
| --- | --- | --- |
| Missing | 4 | 1% |
| Categorised as 'at risk' group<sup>1</sup> |  |  |
| Yes | 72 | 13% |
| No | 486 | 87% |
| Missing | 2 | 0% |
| Received mental health treatment before the pandemic |  |  |
| Yes | 97 | 17% |
| No | 456 | 81% |
| Missing | 7 | 1% |
| Received mental health treatment during the pandemic lockdown |  |  |
| Yes | 34 | 6% |
| No | 518 | 93% |
| Missing | 7 | 1% |
| Direct Covid-19 experience | 39 | 7% |
| Tested positive for Covid-19 | 7 | 1% |
| Experienced symptoms of Covid-19, not tested | 32 | 6% |
| Missing | 0 | 0% |
| Indirect Covid-19 experience<sup>2</sup> | 110 | 20% |
| Close friend tested positive for Covid-19 | 46 | 9% |
| Family member tested positive for Covid-19 | 32 | 6% |
| Knew someone who died of Covid-19 | 44 | 8% |
| Missing | 0 | 0% |
| Employment status affected by Covid-19 | 84 | 15% |
| Fired from job | 15 | 3% |
| Short-time working | 69 | 12% |
| Missing | 6 | 1% |
| Critical worker |  |  |
| Yes | 214 | 38% |
| No | 330 | 59% |
| Missing | 16 | 3% |
| OxCAP-MH score (mean, SD) | 74.1 | 12.3 |
| Missing | 0 | 0% |
| HADS-D (mean, SD) | 4.72 | 4.09 |
| HADS depression score normal (0-7) | 436 | 78% |
| HADS depression score borderline (8-10) | 62 | 11% |
| HADS depression score abnormal (11-21) | 61 | 11% |
| Missing | 1 | 0% |
| HADS-A (mean, SD) | 6.26 | 4.19 |
| HADS anxiety score normal (0-7) | 362 | 65% |
| HADS anxiety score borderline (8-10) | 108 | 19% |
| HADS anxiety score abnormal (11-21) | 89 | 16% |
| Missing | 1 | 0% |
| MSPSS score (mean, SD) |  |  |
| MSPSS high support (5.1-7) | 416 | 74% |
| MSPSS moderate support (3-5) | 112 | 20% |
| MSPSS low support (0-2.99) | 22 | 4% |
| Missing | 10 | 2% |
| WHO-5 score (mean, SD) | 15.1 | 4.8 |
| Missing | 10 | 2% |

Note:
<sup>1</sup> Participants were categorised as 'at risk' group if they were aged 65 or more, and/or they self-reported at least one of the listed diseases: heart/cardiovascular disease, stroke/cerebrovascular disease, lung disease (e.g. asthma, cystic fibrosis, COPD), liver disease (e.g. hepatitis), cancer;
<sup>2</sup> Respondents included in "direct Covid-19 experience" variable were excluded from this group

### Vulnerabilities

A total of 13% of the respondents (n=72) were categorised as belonging to the ‘at risk’ group based on age and/or co-existing physical health conditions. While 17% of the participants (n=97) reported that they received treatment for mental disorders before the period of interest, only 6% of the participants (n=34) reported receiving mental health treatment during the pandemic. Overall, only 30% of those with a mental health service use history (n=29) reported receiving treatment also during the lockdown. A total of 1% of participants (n=7) had been diagnosed with Covid-19, another 6% (n=32) of the participants experienced Covid-19-like symptoms without being tested, and 20% of the respondents (n=110) had indirect Covid-19 experience through an infected friend and/or family member, or knew someone who died of Covid-19. Employment status was affected for 15% (n=84) of participants (job terminated: 3%, n=15; short-term work: 12%, n=69), and 38% of the respondents (n=214) reported having a job categorised as ‘critical worker’ (Table 1).

The level of missing values for the standardised outcome instruments was low with a maximum of ten observations missing (1.8%) for the outcome measures MSPSS and WHO-5. Mean OxCAP-MH score was 74.1 (SD=12.3). Mean WHO-5 score was 15.1 (SD=4.8) with 31% (n=174) of the respondents reporting a score below 13 indicating low mental well-being [37]. The mean scores on HADS-A and HADS-D subscales were 6.3 (SD=4.2) and 4.7 (SD=4.1), respectively, indicating that respondents on average reported higher levels of anxiety than depression symptoms. A total of 74% of participants (n=416) reached the threshold of >5 for high social support on the MPSS scale. Average scores for the MSPSS subscales were 5.41 for family support, 5.53 for support from friends and 5.96 for support from significant others.

### Correlations between capability well-being, mental health/well-being and social support outcomes

Capability well-being (OxCAP-MH) was signficantly strongly/moderately associated with all other outcome measures, the strongest correlation being with depression (HADS-D: r=-0.64, HADS-A: r=- 0.56, WHO-5: r=0.58, MSPSS: r=0.42). In terms of social support, capabilities and depression had the same strenght of correlations, but of opposite directions. (Table 2).

**Table 2.** Correlations between capability well-being, mental health/well-being and social support outcomes

|  | OxCAP-MH | HADS-D | HADS-A | MSPSS | WHO-5 |
| --- | --- | --- | --- | --- | --- |
| OxCAP-MH | 1 |  |  |  |  |
| HADS-D | -0.64*** | 1 |  |  |  |
| HADS-A | -0.56*** | 0.75*** | 1 |  |  |
| MSPSS | 0.42*** | -0.42*** | -0.30*** | 1 |  |
| WHO-5 | 0.58*** | -0.70*** | -0.67*** | 0.34*** | 1 |

Note: \* $p < 0.05$ , \*\* $p < 0.01$ , \*\*\* $p < 0.001$ ; OxCAP-MH - Oxford CAPabilities questionnaire-Mental Health; HADS-D - Hospital Anxiety and Depression Scale Depression subscale; HADS-A - Hospital Anxiety and Depression Scale Anxiety subscale; MSPSS - Multidimensional Scale of Perceived Social Support; WHO-5 - World Health Organisation-Five Well-being Index

### Outcome associations with different types of vulnerability

Outcome associations with different types of vulnerabilities adjusted for sociodemographics are shown in Table 3. Past mental health treatment had a significant negative effect across all outcome measures with an associated capability well-being score reduction of -6.54 (95%CI: -9.26, -3.82), while direct Covid-19 experience had the second most detrimental impact with an associated capability well-being score reduction of -4.58 (95%CI: -8.54, -0.62). Capabilities were similarly negatively affected also for those who belonged to the category ‘at risk’ (−4.45, 95%CI: -7.68, -1.21). These correspond to capability deprivations of -9% and -6%, respectively, when compared to the average capability level of the study cohort.

**Table 3.** Associations between capability well-being, depression, anxiety, social support, mental well-being and different types of vulnerabilities

| Vulnerabilities | OxCAP-MH score |  | HADS-D score |  | HADS-A score |  | MSPSS score |  | WHO-5 score |  |
| --- | --- | --- | --- | --- | --- | --- | --- | --- | --- | --- |
| | $\beta$ (SE) | 95% CI | $\beta$ (SE) | 95% CI | $\beta$ (SE) | 95% CI | $\beta$ (SE) | 95% CI | $\beta$ (SE) | 95% CI |
| At risk group | **-4.45\*\***<br>(1.65) | -7.68, -1.21 | 0.29<br>(0.56) | -0.80, 1.40 | 0.82<br>(0.57) | -0.30, 1.93 | 0.03<br>(0.17) | -0.31, 0.38 | -0.80<br>(0.66) | -2.09, 0.50 |
| Past mental health treatment | **-6.54\*\*\***<br>(1.38) | -9.26, -3.82 | **1.97\*\*\***<br>(0.47) | 1.05, 2.90 | **2.09\*\*\***<br>(0.48) | 1.16, 3.03 | **-0.44\*\***<br>(0.15) | -0.74, -0.15 | **-1.61\*\***<br>(0.56) | -2.71, -0.50 |
| Direct Covid-19 experience | **-4.58\***<br>(2.01) | -8.54, -0.62 | 1.23<br>(0.69) | -0.12, 2.58 | **1.53\***<br>(0.69) | 0.18, 2.89 | -0.09<br>(0.22) | -0.49, 0.35 | **-1.59\***<br>(0.82) | -3.20, 0.00 |
| Indirect Covid-19 experience | -0.11<br>(1.29) | -2.65, 2.43 | 0.25<br>(0.44) | -0.61, 1.12 | **0.97\***<br>(0.45) | 0.09, 1.85 | **-0.31\***<br>(0.14) | -0.59, -0.04 | 0.04<br>(0.53) | -0.99, 1.08 |
| Employment status affected | -2.31<br>(1.55) | -5.36, 0.74 | 0.80<br>(0.53) | -0.24, 1.84 | 0.81<br>(0.53) | -0.24, 1.90 | 0.01<br>(0.16) | -0.33, 0.31 | -1.20<br>(0.62) | -2.43, 0.03 |
| Critical worker | 0.08 (1.12) | -2.13, 2.30 | -0.33<br>(0.38) | -1.08, 0.43 | -0.32<br>(0.39) | -1.09, 0.44 | -0.07<br>(0.12) | -0.30, 0.17 | -0.49<br>(0.45) | -1.38, 0.40 |
| Constant | 68.90***<br>(4.83) | 59.41, 78.38 | 3.08<br>(1.65) | -1.15, 6.32 | 6.44***<br>(1.66) | 3.17, 9.71 | 6.11***<br>(0.51) | 5.11, 7.12 | 16.35***<br>(1.93) | 12.55, 20.15 |

Note: Standard errors (SE) in parentheses \* $p < 0.05$ , \*\* $p < 0.01$ , \*\*\* $p < 0.001$ ; Statistically significant coefficients ( $p < 0.05$ ) in bold; OxCAP-MH - Oxford CAPabilities questionnaire-Mental Health; HADS-D - Hospital Anxiety and Depression Scale Depression subscale; HADS-A - Hospital Anxiety and Depression Scale Anxiety subscale; MSPSS - Multidimensional Scale of Perceived Social Support; WHO-5 - World Health Organisation-Five Well-being Index

Having employment status affected by the pandemic produced consistently lower capability and mental well-being scores as well as higher depression and anxiety scores, but these associations did not reach statistical significance. We could not observe any significant impacts for the category ‘critical worker’ either.

### Associations between capability well-being and current depression, anxiety and social support levels

Additional associations between current levels of depression and anxiety as well as social support with capability well-being were investigated in a separate multivariable regression analysis adjusted for all vulnerabilities and sociodemographics (Table 4). Current levels of depression and anxiety separately showed a capability score reduction of -1.79 (95%CI: -1.99,-1.59) and -1.50 (95%CI: -1.71,-1.29), respectively, per one point difference in the relevant HADS scores. Social support on the other hand proved to be a major capability resilience factor. One point score improvement on the MSPSS scale was associated with an improvement of +3.77 (95%CI: 3.02, 4.53) in the capability scores.

**Table 4.** Associations between capability well-being and current depression (HADS-D), anxiety (HADS- A) and social support (MSPSS) levels

|  | OxCAP-MH |  |
| --- | --- | --- |
| | $\beta$ (SE) | 95% CI |
| HADS-D | -1.76 (0.10)*** | -1.97, -1.56 |
| HADS-A | -1.48 (0.05)*** | -1.70, -1.27 |
| MSPSS | 3.67 (0.38)*** | 2.92, 4.45 |

Note: \* $p < 0.05$ , \*\* $p < 0.01$ , \*\*\* $p < 0.001$ ; Analyses adjusted for age, gender, migration status, education level, children, employment, at risk group for Covid-19, employment status affected by Covid-19, past mental health treatment; OxCAP-MH - Oxford CAPabilities questionnaire-Mental Health; HADS-D - Hospital Anxiety and Depression Scale Depression subscale; HADS-A - Hospital Anxiety and Depression Scale Anxiety subscale; MSPSS - Multidimensional Scale of Perceived Social Support; WHO-5 - World Health Organisation-Five Well-being Index

## Discussion

This is the first study to assess the impact of the Covid-19 lockdown on capabilities in association with mental health/well-being, social support and with different specific vulnerabilities as observed in Austria. Our findings that Covid-19 direct experience is associated with intensified anxiety symptoms, lower mental well-being and lower capabilities is in line with other recent studies exploring the impact of the Covid-19 pandemic on mental health and well-being in Austria [28, 29, 45-47]. Our study showed that participants who reported mental health treatment before the Covid-19 pandemic reported worse outcomes on all measures, including the OxCAP-MH, HADS-D, HADS-A, MSPSS and WHO-5. However, only the OxCAP-MH capability questionnaire showed a significant negative impact for participants categorised as belonging to the ‘at risk’ group. This association has not been captured by any other outcome measure, suggesting an increased sensitivity of the OxCAP-MH in comparison to the other scales used in this study and confirming the advantage of its broader measurement scope when assessing the well-being impact of a pandemic and related public health measures. The study also confirmed that the capability approach, which provides an indication of people’s freedom to engage in forms of being and doing that are of intrinsic value to the person, has direct relevance to situations/policies that inherently limit personal freedoms, i.e. public health emergencies.

The vulnerabilities referred to in this study as ‘employment status affected’ by Covid-19 or being a ‘critical worker’ were not significantly associated with any of the outcomes. Besides the issue of sample size, it may also reflect the Austrian government’s employment support policy implemented in the early stages of the pandemic including the introduction of the short-term working scheme to help retain jobs [48, 49].

When considering the average capability well-being score observed in our cohort, the relative impact of different vulnerabilities and other factors on capability levels were estimated between -9% for those reporting past mental health treatment vs. +5% for reporting one score higher on the social support scale. In future analyses, the outcome scores obtained in this study could also be compared to scores observed in studies prior to the Covid-19 pandemic to further asses the overall impact of this public health emergency and lockdown on the well-being of the Austrian population. Previous studies using the WHO-5 instrument found that 26-27% of the Austrian sample reported scores corresponding to low mental well-being [50, 51]. This is lower than the 31% of respondents who were identified as having low mental well-being (WHO-5 score below 13) in our study. Furthermore, 19% of the participants in this study had borderline and 16% ‘abnormal’ anxiety levels according to HADS-A scoring system, somewhat higher than the levels reported in earlier Austrian studies [52-55]. These results seem to be confirmative of the expected negative impacts of the Covid-19 pandemic, including those of the lockdown, on mental well-being including increased levels of anxiety and stress. Previous studies using the MSPSS scale in Austrian populations reported comparable scores, indicating relatively high social support [56, 57].

In addition to providing an indication of the Covid-19 and lockdown impacts on vulnerable groups, this study also presents the interactions between capability well-being levels and current mental health indicating a strong negative impact of current depression and anxiety. On the other hand, social support was shown as a major capability resilience factor. Future (public health) policies should take the strong associations between capabilities and current mental health and social support levels directly into consideration to minimise the negative long-term health, social and economic issues related to future public health emergencies.

Furthermore, our results suggest that amongst all investigated vulnerabilities, people with past mental health treatment represent the most vulnerable group. A recent study from Austria found that the number of people treated with psychotherapy during lockdown (personal, phone or virtual contacts) decreased by one-third [58]. In our study, the proportion of people receiving mental health treatment during lockdown in comparison to the period before the pandemic was 6% vs. 17%, respectively. We found indication for treatment continuation between the two periods for only 30% of those participants who received mental health treatment prior to the pandemic. Even under the most conservative assumptions, these results suggest a substantial level of underutilisation of mental health services (due to whatever causes) during the lockdown period. For future strategic healthcare planning during next waves of the pandemic, policy makers and health and social care providers need to be aware of the exceptional vulnerability of this group and efforts have to be made for continuous mental health service provision. Digital e-health treatment options provide potential solution to assure the continuity of treatment and at the same time protect health of the service-users and professionals [59, 60].

The main limitation of our study is that the participants completed the survey retrospectively about one month after the lockdown (mid-May 2020). This time lag may have introduced some recall bias considering the self-reported outcome measures. Since data were collected at the time when the number of new Covid-19 cases were relatively low and the Austrian epidemic curve has flattened, we assume that the presented estimates are more conservative and optimistic than if the survey questions would have been completed directly during the lockdown. Moreover, since the analysis is based on one measurement point, the study allows no causal conclusions. Our study is also prone to limitations of online survey; results are based fully on self-reporting with the potential to reporting bias [61] and some groups (females, younger ages, higher educated), were over-represented in the survey sample compared to the general population [62, 63]. The survey on the other hand achieved satisfactory representation in terms of more than half of the Austrian provinces, migration background and employment status.

## Conclusions

This research contributes to the understanding of the impact that pandemics and nationwide responses to pandemics can have on mental health and broader capability well-beings in light of their major policy relevance. Furthermore, the study confirms that the OxCAP-MH capability measure is a valid and relevant tool to understand the impacts of the Covid-19 pandemic and related public health measures which due to the negative externalities of any infectious disease inherently limit individual freedoms to some extent. Future research is planned to compare cultural aspects of lockdown experiences across countries and explore long-term mental health/well-being impacts from the perspective of the capability approach.

## Data Availability

The dataset generated during the current study is available from the corresponding author on a reasonable request.

## List of abbreviations

HADS: The Hospital Anxiety and Depression Scale
MSPSS: The Multidimensional Scale of Perceived Social Support
OxCAP-MH: The Oxford CAPabilities questionnaire-Mental Health
WHO: World Health Organisation
WHO-5: The World Health Organisation-Five Well-being Index

## Declarations

### Ethical approval and consent to participate

All procedures performed in studies involving human participants were in accordance with the ethical standards of the Ethics Commission of the Medical University of Vienna (EK 1529/2020) and with the 1964 Helsinki declaration and its later amendments or comparable ethical standards. Informed consent was obtained from all individual participants included in the study.

### Consent for publication

Not applicable.

### Availability of data and materials

The dataset generated during the current study and the study protocol have been released in a scientific data repository and can be accessed under the link: https://doi.org/10.5281/zenodo.4271534.

### Authors’ contributions

JS, RW and CV conceived the study idea, and developed the conceptual framework and methods of the research. JS provided the resources to this study. TH and AL executed the survey and conducted the analysis supervised by JS. JS, TH and AL wrote the manuscript which was reviewed by all. All authors provided critical feedback and helped shape the research, analysis and manuscript. All authors approved the final manuscript.

## Acknowledgements

We would like to say thank you to all survey participants and to colleagues at the Department of Health Economics for piloting the survey.

